# Sourcing Bivariate Genetic Overlap for Polygenic Prediction using MiXeR-Pred

**DOI:** 10.1101/2024.02.19.24303039

**Authors:** Nadine Parker, Weiqiu Cheng, Guy F. L. Hindley, Kevin S. O’Connell, Pravesh Parekh, Espen Hagen, Olav B. Smeland, Srdjan Djurovic, Alexey A. Shadrin, Ole A. Andreassen, Oleksandr Frei, Anders M. Dale

## Abstract

The past two decades have seen the advent and mass application of genome-wide association studies (GWAS). The observation that complex phenotypes are polygenic has contributed to the development of the polygenic score (PGS) for understanding individual-level genetic predisposition. There have been substantial advances in PGS methodology in recent years. However, few methods leverage the pleiotropic nature of complex phenotypes for polygenic prediction. Here, we present MiXeR-Pred, a novel approach for polygenic prediction that builds on an established MiXeR framework to source genetic overlap with a secondary phenotype to inform PGS prediction of a primary phenotype. We apply MiXeR-Pred using both bipolar disorder and schizophrenia as complex primary phenotypes along with the following secondary phenotypes: education attainment, major depressive disorder, and measures of cortical brain morphology. We compare MiXeR-Pred predictions to the PGS derived from each primary phenotype’s GWAS in addition to the multi-trait analysis of GWAS (MTAG) approach, which can use correlated secondary phenotypes to boost discovery and prediction for a primary phenotype. We show that MiXeR-Pred improves prediction performance when compared to both the primary GWAS and MTAG PGS, regardless of the secondary phenotype. Not only can MiXeR-Pred be used to further our understanding of pleiotropy among complex phenotypes, but it also provides a novel conceptualization of how one can source pleiotropy to improve PGS performance which can ultimately contribute to advancements in personalized medicine. The MiXeR-Pred tool is available at https://github.com/precimed/mixer-pred.

## Introduction

The past 20 years have seen the advent and mass application of genome wide association studies (GWAS) performed for a wide range of phenotypes. These studies have revealed that complex phenotypes are polygenic with many associated genetic variants having small effect sizes.^1^ The polygenic nature of a complex phenotype can be leveraged to compute a polygenic score (PGS). Representing an individual’s genetic predisposition for a given phenotype, a PGS is typically computed as a weighted sum of an individual’s allele count across genetic variants, with weights derived from a corresponding GWAS.^1,2^ While the PGS approach has widespread utility there is still a need for improvement with respect to complex human diseases.^3,4^

For many complex phenotypes, there remains large differences in the phenotypic variance explained by their PGS and the estimated heritability based on associated single nucleotide polymorphisms (i.e., SNP-based heritability). Methodological advancements in computing PGSs have resulted in improved prediction.^5,6^ Many of the recent PGS methods attempt to move from effects estimated in GWAS for SNPs tagging particular genomic loci to accurate modeling of “direct causal effects” of a given SNP on a phenotype refined from the effects of other SNPs propagated through linkage disequilibrium (LD).^7–9^ However, the development of differing strategies that leverage the accumulating knowledge of complex phenotype genetics can contribute to prediction performance for these various traits and disorders.

The accumulated GWAS findings have revealed that there is widespread pleiotropy across many complex phenotypes.^10^ This pleiotropy can be observed through genetic correlations or other approaches that measure genetic overlap beyond genetic correlations by estimating the number of overlapping loci or genetic variants.^11–13^ Pleiotropy has been leveraged to improve genetic discovery due to the increase in statistical power gained by combining signal from genetically related phenotypes.^14,15^ This concept has also been extended to PGS methods, where genetic overlap between phenotypes has been leveraged to improve the prediction of a primary GWAS phenotype.^14,16^ One approach uses multiple polygenic scores for the prediction of a single phenotype.^17–19^ However, this approach doesn’t produce a single, improved polygenic score for the primary phenotype of interest. Comparatively, the multi-trait analysis of GWAS (MTAG) approach can use genetically correlated secondary phenotypes to boost discovery and prediction for a primary phenotype.^14^ Moreover, MTAG utilizes standardized effect estimates (i.e., Z-scores) which is also exploited in the standard meta-analytic GWAS approach.^20^ The use of Z-scores may denoise genetic signal from GWAS and potentially improve prediction. Still, approaches that leverage genetic overlap for prediction are lacking. Moreover, identifying the effect of pleiotropy on polygenic prediction can advance our understanding of the genetically overlapping phenotypes themselves.

We have developed a Gaussian causal mixture model (MiXeR) framework which is the basis for a growing suite of tools. For a pair of phenotypes, MiXeR estimates genetic overlap by modeling SNPs with a non-zero effect on the phenotype (i.e., non-null variants) and those with no effect (i.e., null variants) and approximates the distribution of shared and phenotype-specific non-null variants.^12^ Notably, this bivariate approach can produce a non-correlative estimate of genetic overlap. Here, we extend the MiXeR framework to include MiXeR-Pred, a tool for polygenic prediction. MiXeR-Pred predicts a primary phenotype sourcing genetic overlap with a secondary phenotype. To illustrate the utility of MiXeR-Pred, we use bipolar disorder (BIP) and schizophrenia (SCZ) as our primary phenotypes. BIP and SCZ are complex phenotypes with known genetic overlap and PGSs that explain only a portion of SNP-based heritability.^21–23^ Moreover, we source pleiotropy from several secondary phenotypes including education attainment (EDU), major depressive disorder (MDD), and cortical brain morphology that exhibit correlative and non-correlative genetic overlap with BIP and SCZ.^21,24–30^ We demonstrate that MiXeR-Pred improves prediction of both BIP and SCZ when compared to PGS derived from the primary GWAS only and PGS derived from the MTAG approach.

## Methods

MiXeR-Pred is an extension of previous MiXeR implementations based on Gaussian causal mixture models.^12,31^ In the univariate setting, MiXeR models non-null and null SNPs using spike-and-slab prior distributions, and accounting for LD. MiXeR then estimates the proportion of non-null SNPs (i.e., polygenicity) and the variance of non-null effects (i.e., discoverability). The bivariate MiXeR extension estimates the distribution of shared and phenotype-specific non-null SNPs between a pair of phenotypes. The most recent gene set analysis version of MiXeR (GSA-MiXeR) expands the univariate MiXeR model to incorporate functional annotations, thus more accurately modeling genetic architecture of complex phenotypes.^31^ MiXeR-Pred extends previous bivariate MiXeR models to include the functional annotations modelled in GSA-MiXeR, which has been shown to improve prediction and GWAS replication rates.^32–34^ In addition, MiXeR-Pred expands on the standard pruning and thresholding model by modifying both selection of the SNPs to be included in the PGS calculation and their respective weights. These parameters are derived from the posterior estimates of SNP effect sizes conditioned on GWAS Z-scores of both the primary and secondary phenotypes.

Overall, MiXeR-Pred utilizes information on SNP functional annotations, heterozygosity, LD, and pleiotropy to improve prediction of a primary phenotype using a secondary phenotype.

### MiXeR-Pred Parameter Estimation

MiXeR models each SNP, i, with an additive genetic effect, β_I_, as a point normal mixture model 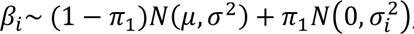, where π_1_ is the prior probability of a SNP having a non-zero effect (polygenicity) while σ_i_^2^ gives the effect size variance (discoverability) and is allowed to vary across SNPs, and *N*(μ, σ^2^) denotes a Gaussian distribution with mean μ and variance σ^2^. In line with our new GSA-MiXeR^31^ models, the number of effective parameters being optimized is reduced by parametrizing 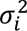 based on functional categories, enrichment in coding genes, allele frequency, and LD score as follows:

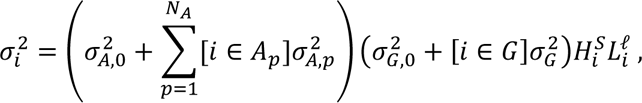

where the 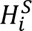 term allows for incorporation of allele frequency on genetic architecture, the term *H*_*i*_ = 2*f*_*i*_ (1 − *f*_*i*_) denotes heterozygosity of the *i*-th SNP where *f*_*i*_ is the minor allele frequency of the *i*-th SNP, and *S* controls effect size distribution across the spectrum of allele frequencies. LD-dependent genetic architectures are modelled with the 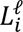 term, where *L*_*i*_ denotes the LD score of the *i*-th SNP and ℓ parameter controls the effect size distribution with respect to LD score. The index *p* runs across functional categories {*A*_1_, *A*_2_,…, *A*_*NA*_}. If a SNP belongs to the *p*-th functional category, as indicated by [*i* ∈ *A*_*p*_] term, 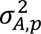 gives the contribution of the functional category to the variance of the *i*-th g SNP. Parameters 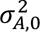 and 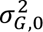 allow for non-zero variance 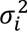 for SNPs that do not belong to any functional category or genes, and genomic region *G* represents a union of all protein-coding genes, allowing the modelling of the overall enrichment in effect sizes in coding genes.

MiXeR inference of parameters is based on maximizing the log-likelihood function log *L*(*z*_1_,…, *z*_*m*_|θ) → max observing a set of GWAS summary statistics given model parameters, while accounting for the LD structure among SNPs and their allele frequencies. Details on likelihood computation and optimization have been described elsewhere.^31^ Here, MiXeR-Pred uses allele frequencies and LD structure estimated from the Haplotype Reference Consortia (HRC) reference panel with 11,980,511 SNPs and 23,152 samples after applying the basic QC procedure described by Frei et al. (2023).^31^

The bivariate MiXeR analysis models additive genetic effects as a mixture of four components: (1) SNPs that are null in both phenotypes (π_0_), (2) SNPs uniquely affecting the first phenotype π_1_, (3) SNPs uniquely affecting the second phenotype (π_2_), and (4) SNPs with an effect on both phenotypes (π_12_).^12^ In the shared component, bivariate MiXeR models the variance-covariance matrix as 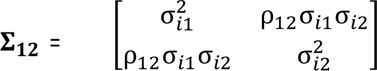 where ρ_12_ denotes the genetic correlation (r_g_) within the shared component, and 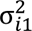 and 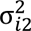 are the discoverability parameter estimated in the univariate analysis of the two phenotypes.

In the MiXeR model, GWAS Z-scores of j-th SNP in two phenotypes are modelled as follows:

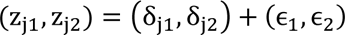

Where (δ_j1_, δ_j2_) is the true genetic component, and 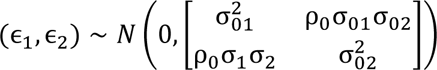 is the residual variation. Applying Bayesian rules to this model one can derive the posterior distribution of the true genetic component given observed GWAS Z-scores and estimated parameters θ of the 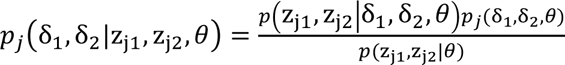

MiXeR-pred utilizes the first *E*δ_j1_ and the second 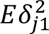 moments of the posterior distribution:

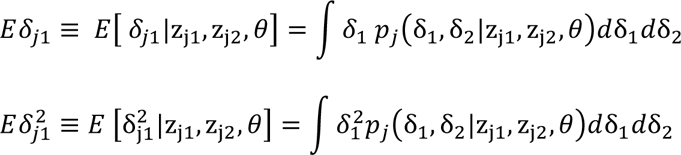

The first moments are used as weights for MiXeR-Pred and the second moments are used to inform the selection of SNPs into the model.

We also, generate a univariate MiXeR-Pred models for comparison with the main bivariate models. Univariate MiXeR-pred similarly utilizes the first *E*δ_j1_ and the second 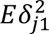 moments of the posterior distribution based on the primary phenotype only:

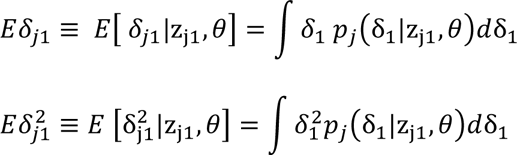

### MiXeR-Pred Polygenic Score Calculations

MiXeR-Pred implements the pruning and thresholding approach to PGS using *E*δ_1_as SNP weights and the exponentiated negative value of 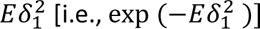 as thresholds for SNP selection. This transformation of 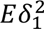 ensures the values range between 0 and 1, with stronger associations having a lower value and serves a similar purpose as p-values in the standard pruning and thresholding approach.

After the weights and thresholds are generated, there are several minor processing steps. First, strand-ambiguous SNPs were removed. The remaining SNPs were filtered to be among those present in the target sample. Next, we use PLINK v1.9^35^ for clumping, generating a list of independent genetic SNPs (details in the Supplementary Methods). These independent SNPs are then used to construct a PGS with MiXeR-Pred weights using PRSice2.^36^

### Samples

We conduct our PGS analysis using the Norwegian Thematically Organized Psychosis (TOP) sample. Briefly, a total of 440 indiviudals with bipolar disorder [BIP; mean age (sd)=34.72 (12.84), 60.68% female], 696 individuals with schizophrenia [SCZ; mean age (sd)=32.90 (13.37), 42.53% female], and 1044 controls [mean age (sd)=32.51 (9.96), 47.51% female] of European ancestry were included in this study. More details on the TOP sample and basic quality control of genetic data can be found in the Supplementary Methods.

### GWAS Summary Statistics

We acquired GWAS summary statistics for both BIP and SCZ as well as related phenotypes with both correlated and uncorrelated genetic effects: education attainment (EDU), major depressive disorder (MDD), brain cortical thickness (TH) and surface area (SA). We used GWAS summary statistics from the Psychiatric Genomics Consortium, excluding the TOP sample participants, for the BIP GWAS^22^ with 413,466 participants (41,917 cases) and the SCZ GWAS^23^ with 130,644 participants (53,386 cases). For EDU, GWAS summary statistics were acquired from the Social Science Genetic Association Consortium comprising 765,283 participants after excluding the 23andMe sample.^37^ For MDD we used the recent GWAS by Als et al (2023)^38^, with 1,349,887 participants (371,184 cases). For both cortical TH and SA we used GWAS performed on 32,877 UK biobank participants with no prior mental disorder diagnosis.^27^ All GWAS data were restricted to participants of European ancestry.

### Evaluating Prediction Performance

To compare PGS performance we used two main approaches. First, we used PRSice2 to calculate PGS R^2^ (Supplementary Methods). Our main results present the partial R^2^ on the liability scale for the PGS effect after adjusting for the genetic batch and first 20 genetic principal components. The liability scale R^2^ was calculated using the approach by Lee et al. (2012)^39^ with a population prevalence of 2% and 1% for BIP and SCZ, respectively. As a second approach, we split the sample into a 70% training set and 30% out of sample test set. In the 70% training set, we performed 5-fold cross-validation of generalized linear models. These models assessed PGS effect which were pre-residualized for genetic batch and the first 20 genetic principal components. Next, the best performing generalized linear model in the cross-validation procedure was tested on the held-out 30% of the sample (i.e., out of sample test set). The reported prediction performance is the area under the receiver operator curve (AUC) estimated in the 30% out of sample test set. Note, for each primary phenotype, the same sample is used for the training set and out of sample test set throughout all analyses. This procedure was implemented using the caret package in R v4.0.0. In addition, we performed sensitivity analyses adjusting PGS effect for age, sex, genetic batch, and the first 20 genetic principal components using both approaches.

### Comparison of Polygenic Risk Score Weights

Both estimation of SNP weights and SNP selection contribute to PGS performance. MiXeR-Pred weights, and by extension thresholds, are derived from posterior estimates utilizing GWAS Z-scores. Since most PGS methods use beta (or log odds ratio) estimates, we investigate the effect of using Z-scores and inverse variance weighted 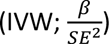 values (i.e., standardized effect estimates) compared to beta estimates as weights for prediction of BIP and SCZ. We used PLINK v1.9 for clumping (Supplementary Methods) and PRSice2 for PGS calculations and PGS R^2^ estimation. In each of the three scenarios (i.e., beta, Z, IVW), the p-value from the phenotype-specific GWAS was used for pruning and thresholding. For comparison across the three weights, 33 PGS were generated using p-value thresholds that included the top 1,000 to 300,000 independent SNPs.

To investigate the relationship between MiXeR-Pred weights and the above primary GWAS weights, we used Pearson correlations between MiXeR-Pred weights and the primary GWAS beta, Z, and IVW values. In addition, Pearson correlations were calculated between MiXeR-Pred thresholding values and the primary GWAS Z, IVW values as well as MiXeR-Pred weights.

### Method Comparisons

We illustrate the performance of MiXeR-Pred by predicting BIP and SCZ (primary phenotypes) using several genetically correlated and uncorrelated secondary phenotypes (EDU, MDD, SA, and TH). Each primary phenotype also served as a secondary phenotype for each other. As our main method comparison, we use MTAG.^14^ For a given phenotype, MTAG can use additional phenotypes to boost discovery and prediction. We also used the PGS for the primary phenotype as an additional comparison. The MTAG outputs and the GWAS summary statistics for each primary phenotype were minimally processed using the same steps as MiXeR-Pred. Ambiguous SNPs were removed and the remaining SNPs were filtered to be among those present in the target sample. PLINK v1.9^35^ was used for clumping with the same parameters as MiXeR-Pred (Supplementary Methods). The independent SNPs identified by clumping were then used to compute polygenic scores using PRSice2.^36^ Here we used Z-scores as weights for MTAG and the GWAS for comparability with MiXeR-Pred weights which are derived from GWAS Z-cores. For both MTAG and the GWAS we tested the predictive performance for 33 PGSs based on p-value thresholds, ranging from including the top 1,000 to 300,000 independent SNPs. The predictive performance of these MTAG and GWAS PGS were comparted to the 33 PGSs derived with MiXeR-Pred weights and thresholds.

### Evaluating Contributions to MiXeR-Pred Prediction

Given that MiXeR-Pred modelling of genetic architecture may contribute to improved prediction, we generated univariate PGS for BIP and SCZ at the same 33 top SNP thresholds. To evaluate the potential contribution of MiXeR-Pred at a univariate level, we compared their prediction performance to that of the bivariate MiXeR-Pred PGS. To assess the potential role of uncorrelated phenotypes with the same overlap in genetic variants estimated by MiXeR-Pred, we took correlated secondary phenotypes and randomly changed effect directions across SNPs. The resulting “shuffled” and uncorrelated secondary phenotypes were then used to generate PGS using MiXeR-Pred. The performance of these PGS were then compared to that of univariate MiXeR-Pred for a given primary phenotype. We also assessed the separate contributions of MiXeR-Pred weights and thresholds for SNP selection at the univariate and bivariate levels (Supplementary Methods).

To determine which aspects of MiXeR modelling contributes to prediction, we compared several bivariate MiXeR parameters (Supplementary Methods) with MiXeR-Pred prediction performance. These parameters from bivariate MiXeR v1.3 include estimates for (i) the degree of non-correlative genetic overlap in estimated shared variants between a primary and secondary phenotype (dice coefficient), (ii) correlated overlap (genome-wide r_g_ and r_g_ within the shared component), and (iii) the fraction of SNP-heritability explained by genome-wide significant variants for the secondary phenotype given the current GWAS sample size (power). We tested for correlation between these parameters and MiXeR-Pred performance metrics (R^2^ and AUC).

## Results

### Comparison of Polygenic Score Weights for Prediction

The effect of using beta, Z, and IVW as PGS weights was evaluated in a sample of n=440 BIP, n=696 SCZ, and n=1044 controls (Figure 1). For BIP, IVW achieved the highest prediction performance (max: liability R^2^=0.05, AUC=0.65) followed closely by using Z as weights (max: liability R^2^=0.05, AUC=0.64). For SCZ, Z weights achieved the highest prediction performance (max: liability R^2^=0.07, AUC=0.68) followed closely by both beta (max: liability R^2^=0.07, AUC=0.68) and IVW (max: liability R^2^=0.07, AUC=0.68) as weights.

**Figure 1.**
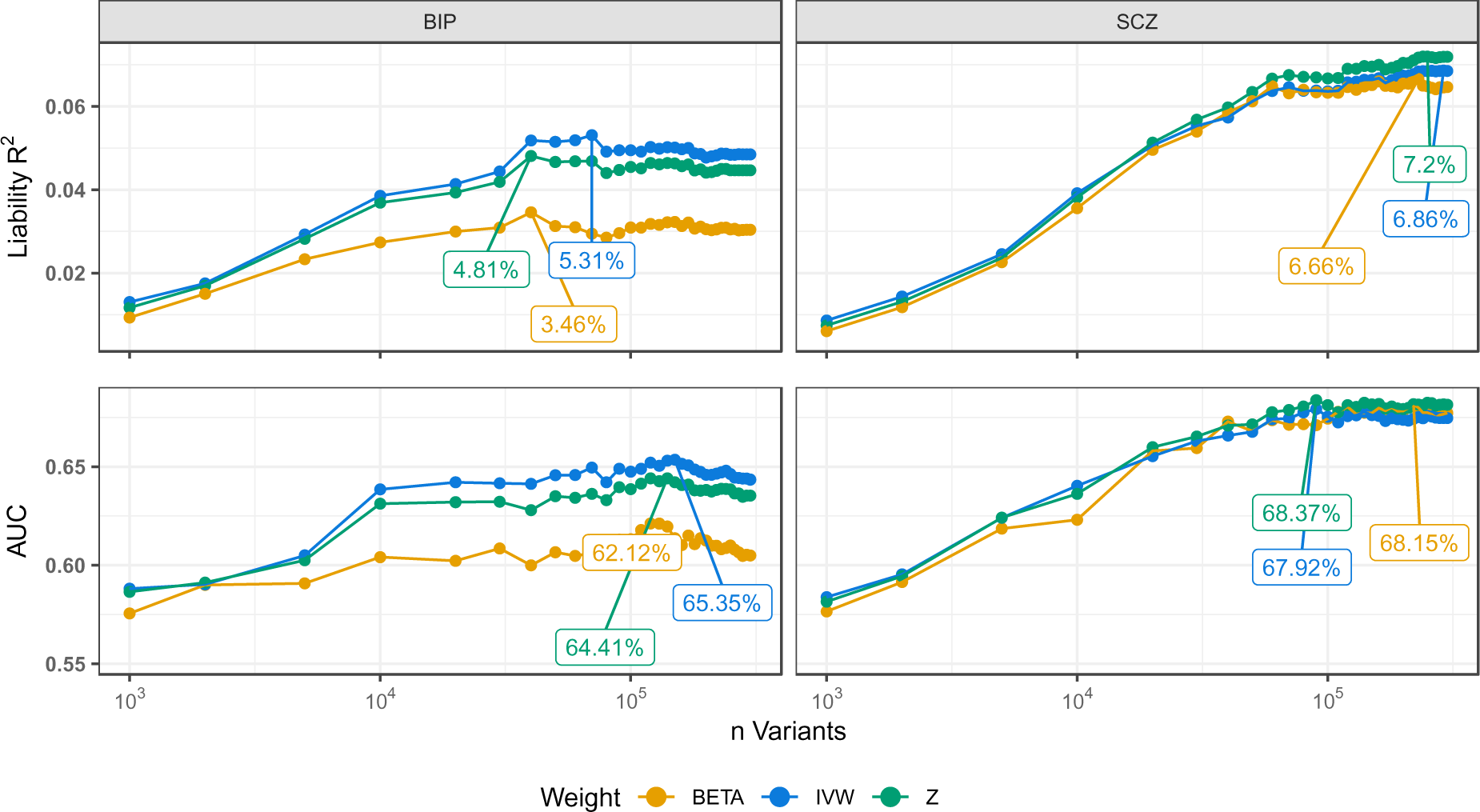
Comparison of Weights for Polygenic Scores. Prediction performance, measured as variance explained (R^2^) on the liability scale (top row of figures) and the area under the receiver operator curve (AUC; bottom row of figures), for polygenic scores (PGS) derived for bipolar disorder (BIP; left) and schizophrenia (SCZ; right) are presented. Population prevalence of 2% and 1% were used to estimate liability R^2^ for BIP and SCZ, respectively. PGS were calculated using thresholds for top variants from 1000 to 300,000, shown on the x-axis using a log10 scale. The three labelled values in each plot represent the maximum prediction performance for each of the three weights expressed as percentages.

For both BIP and SCZ, MiXeR-Pred weights were compared to the primary phenotypes GWAS beta, Z, and IVW. Figure 2 shows that MiXeR-Pred weights (*E*δ_1_) are strongly correlated with the primary phenotypes Z (BIP: r=0.74, p<0.001; SCZ: r=0.83, p<0.001) and IVW (BIP: r=0.84, p<0.001; SCZ: r=0.91, p<0.001) values but substantially less correlated with betas (BIP: r=0.13, p<0.001; SCZ: r=0.14, p<0.001). Meanwhile, MiXeR-Pred thresholding values (exp(−*E*δ_1_^2^)) coincide well with the primary phenotypes Z and IVW values but, as expected, were strongly associated with MiXeR-Pred weights (Figure 2 B & D).

**Figure 2.**
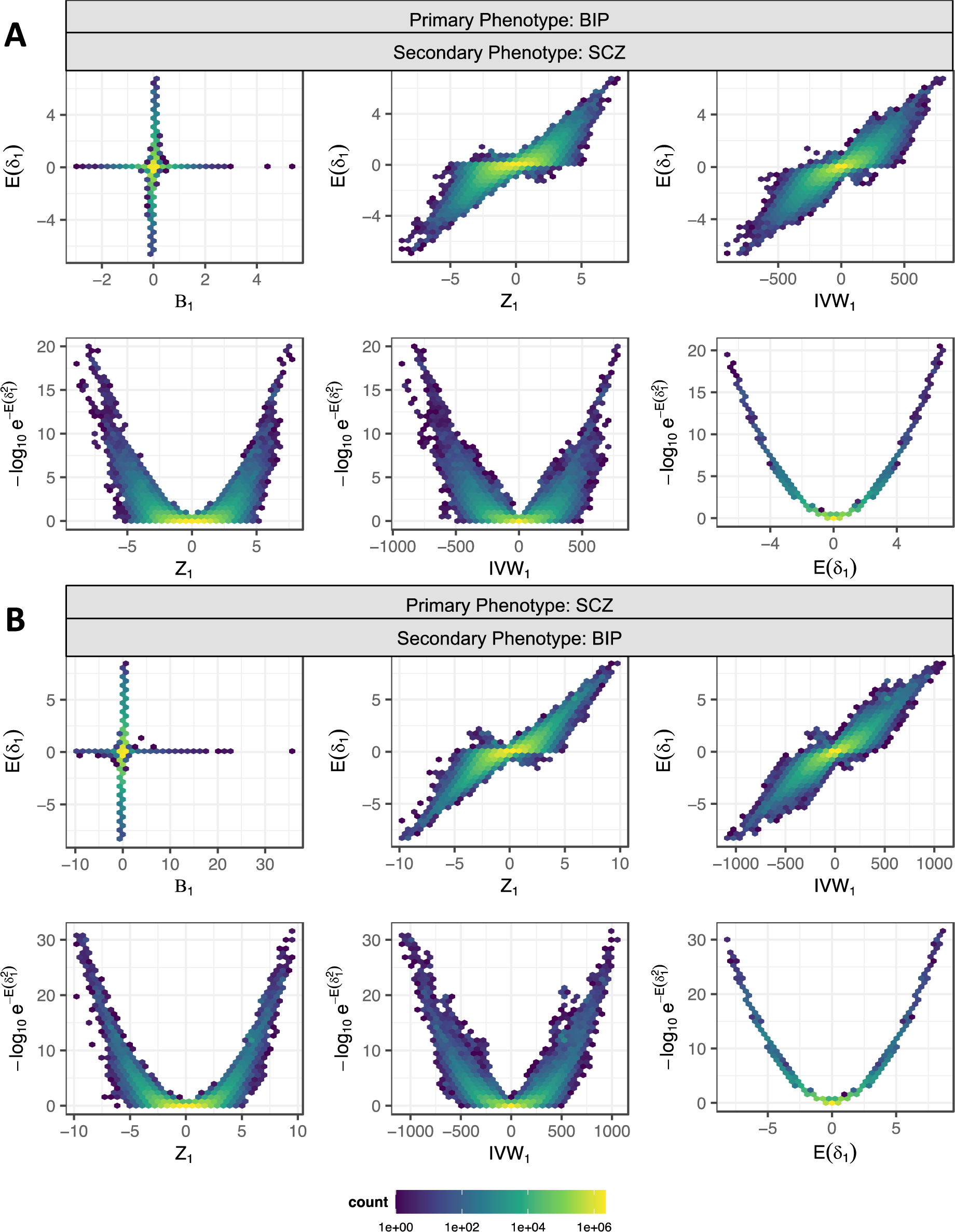
Comparison of MiXeR-Pred Estimates with Primary Phenotype Weights. Density plots for the number of single nucleotide polymorphisms where panel (A) depicts comparisons for bipolar disorder (BIP) as the primary phenotype and schizophrenia (SCZ) as the secondary phenotype. Panel (B) depicts comparisons for SCZ as the primary phenotype and BIP as the secondary phenotype. MiXeR-Pred weights (*E*δ_1_) are compared with the primary phenotype’s beta (B_1_), Z-score (Z_1_), and inverse variance weighted (IVW_1_) values using density plots top row of panels A and B. In the bottow rows, MiXeR-Pred thresholds 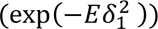 are -log_10_ transformed and compared with the primary phenotype’s Z_1_ and IVW_1_ values as well as MiXeR-Pred weights.

### Improved Prediction Performance with MiXeR-Pred

We compared MiXeR-Pred prediction performance for the two primary phenotypes (BIP and SCZ) across multiple secondary phenotypes that exhibit varying degrees of polygenicity and genetic correlation (Figure 3A; supplementary table 1). Across all secondary phenotypes, MiXeR-Pred exhibited improved prediction (Figure 3B and C; Supplementary Figure 1).

**Figure 3.**
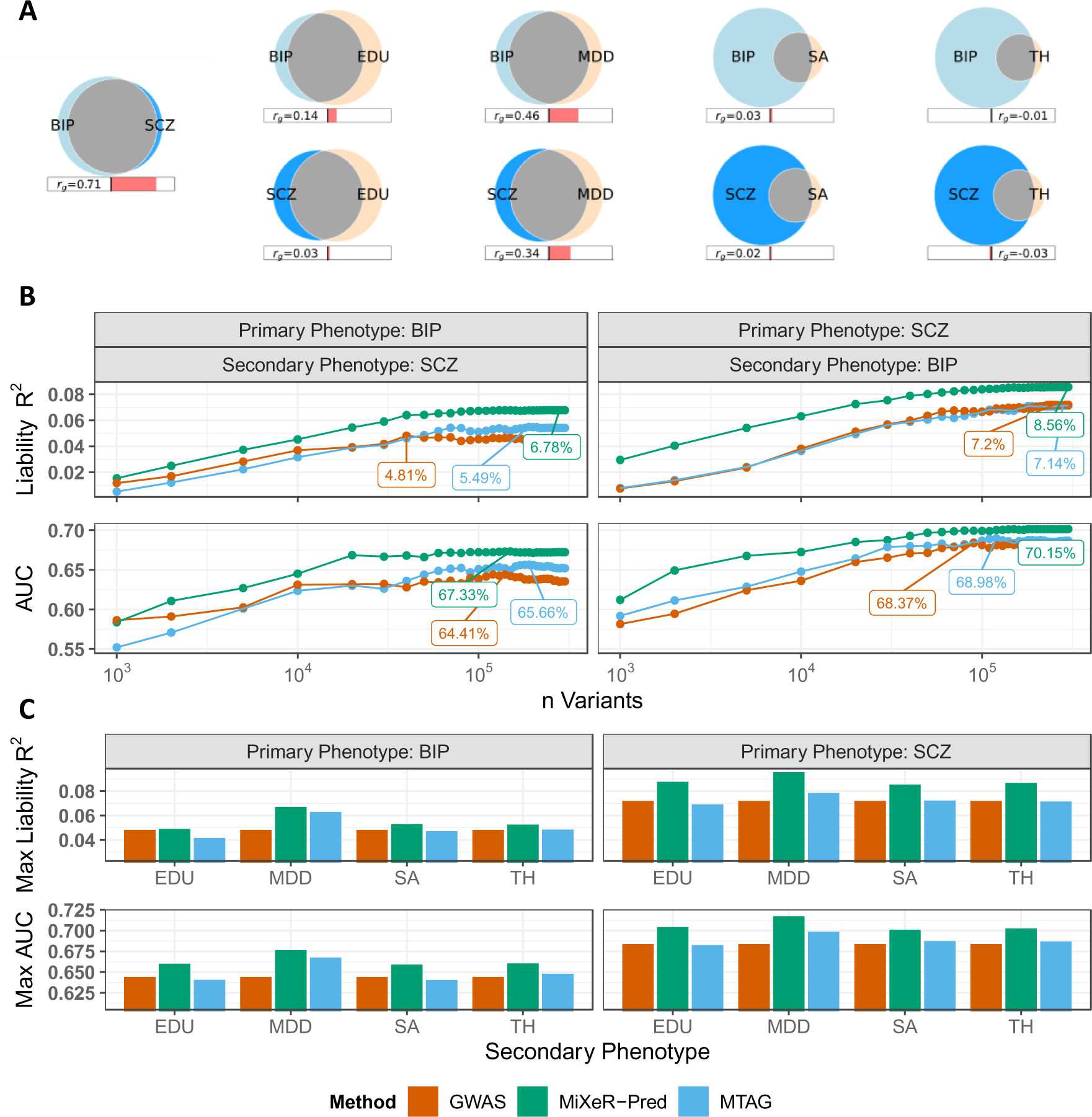
Comparison of Prediction Performance Across Methods and Secondary Phenotypes. (A) Venn diagrams illustrating MiXeR estimates of: (i) the polygenicity of each phenotype (size of the circles) and for each pair of primary and secondary phenotype (ii) the degree of non-correlative genetic overlap (grey shaded overlapping region) and (iii) the genetic correlation (r_g_; value and bar below the Venn diagrams). Panels (B) and (C) compare the MiXeR-Pred polygenic score (PGS) prediction performance [variance explained (R^2^) on the liability scale and area under the receiver operator curve (AUC)] to the multi-trait analysis of genome wide association studies (MTAG) and the primary phenotype’s genome wide association study (GWAS) PGS (using Z-scores as weights). In (B) BIP and SCZ were used as primary and secondary phenotypes for each other. On the x-axis the number of genetic variants selected for each PGS are shown using a log10 scale. In (C) education attainment (EDU), major depressive disorder (MDD), cortical surface area (SA), and cortical thickness (TH) were used as secondary phenotypes for BIP and SCZ. Note that the secondary phenotypes are used for both MiXeR-Pred and MTAG approaches. The GWAS PGS performance is constant regardless of secondary phenotype since it is the PGS derived from the primary phenotypes GWAS only.

The largest boost in MiXeR-Pred PGS prediction for BIP was observed when using SCZ as a secondary phenotype (max liability R^2^=0.07, max AUC=0.67). While using BIP as a secondary phenotype for SCZ resulted in improved prediction, MDD as a secondary phenotype provided the greatest boost in prediction for both MiXeR-Pred (max liability R^2^=0.10, max AUC=0.72) and MTAG (max liability R^2^=0.08, max AUC=0.70) over the SCZ GWAS (max liability R^2^=0.07, max AUC=0.68) PGS. The secondary phenotype that provided the least boost in prediction was EDU for BIP (MiXeR-Pred: max liability R^2^=0.05, max AUC=0.66; MTAG: max liability R^2^=0.04, max AUC=0.64; BIP GWAS: max liability R^2^=0.05, max AUC=0.64) and SA for SCZ (MiXeR-Pred: max liability R^2^=0.09, max AUC=0.70; MTAG: max liability R^2^=0.07, max AUC=0.69; SCZ GWAS: max liability R^2^=0.07, max AUC=0.68). Of note, models adjusting for demographic covariates (i.e., age and sex) in addition to genetic batch and genetic PCs produced similar results (Supplementary Figure 2).

### Contributions to Improved Prediction Performance

To evaluate the potential contribution of improved modelling of the primary phenotype’s genetic architecture by MiXeR-Pred, we generated univariate MiXeR-Pred PGS for BIP and SCZ and compared prediction performance with that of bivariate MiXeR-Pred PGS. The univariate MiXeR-Pred PGS achieved a max R^2^=0.05 and R^2^=0.09 on the liability scale for BIP and SCZ respectively (max: AUC_BIP_=0.66, AUC_SCZ_=0.70; Supplementary Figure 3). When using MDD and SCZ as secondary phenotypes, bivariate MiXeR-Pred prediction of BIP was higher than univariate (Figure 4). When using MDD, and to lesser extent EDU, as secondary phenotypes, bivariate MiXeR-Pred prediction of SCZ was higher than univariate (Figure 4). To further assess if MiXeR-Pred can leverage genetically uncorrelated secondary phenotypes for prediction, we made genetically correlated secondary phenotypes uncorrelated by randomly changing effect directions (Supplementary Table 2). This augmentation resulted in modest increases in the prediction of BIP when using genetically uncorrelated SCZ and MDD as secondary phenotypes with more subtle effects on SCZ as a primary phenotype (Supplementary Figure 4).

**Figure 4.**
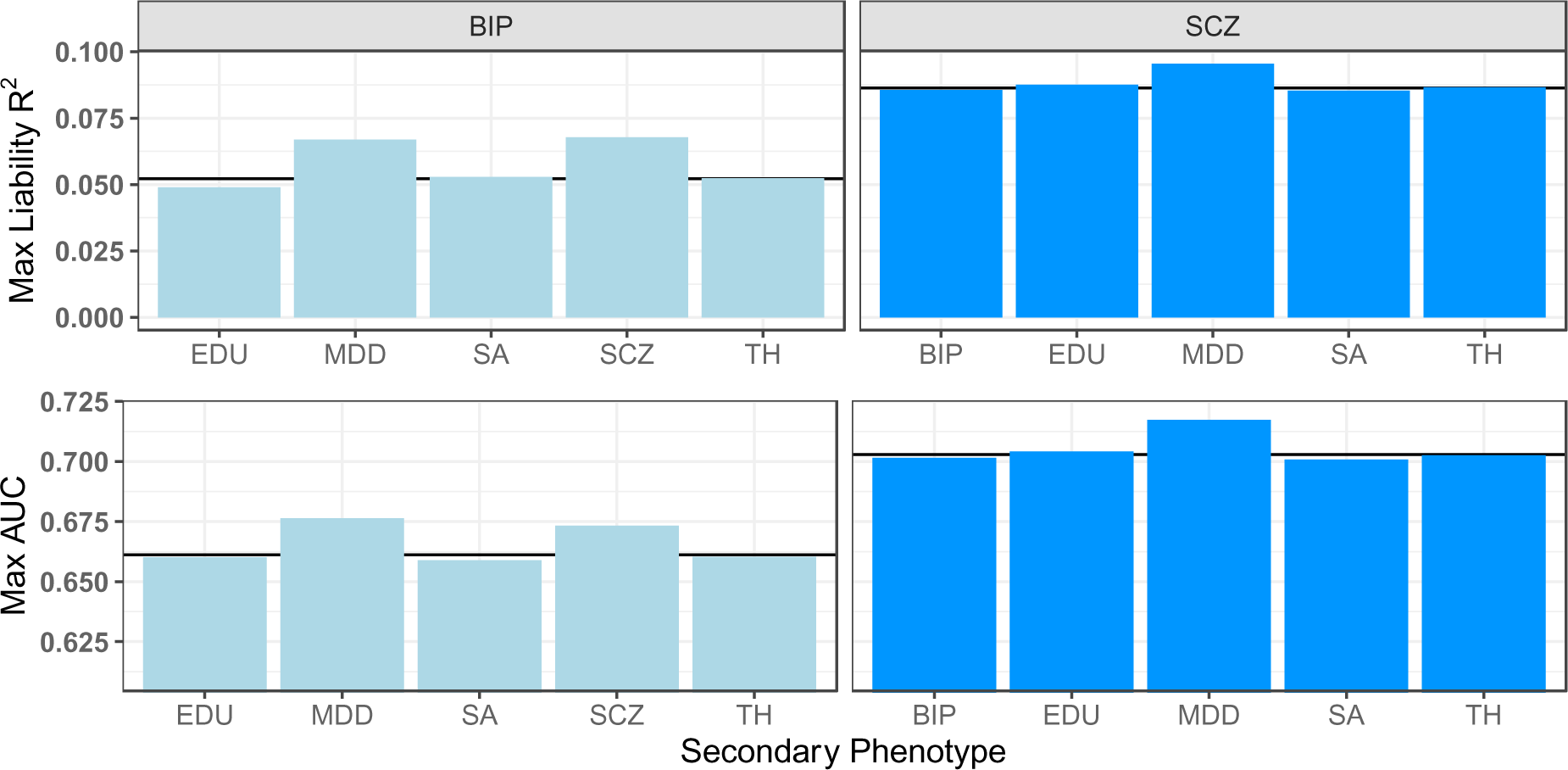
Univariate and Bivariate MiXeR-Pred Prediction Performance. Bar graphs illustrating a comparison of the MiXeR-Pred maximum prediction performance at the univariate level (solid black line) and bivariate level (bars) for the primary phenotypes BIP and SCZ.

We also assessed the separate contributions of MiXeR-Pred weights and thresholds for SNP selection at the univariate and bivariate levels (Supplementary Figures 3, 5, and 6). SNP selection appeared to contribute most to MiXeR-Pred performance as it most often achieved the highest prediction and with fewer SNPs. Notably, the increase in prediction performance associated with SNP selection, compared to selection and weights, was largest for genetically uncorrelated secondary phenotypes (e.g., SA and TH).

To evaluate the effect of various MiXeR estimates on mixer-pred performance, we tested their correlation for each primary phenotype across secondary phenotypes (Figure 5). A significant correlation may indicate that a given mixer estimate may contribute to improved prediction performance. Both the genetic correlation within the shared component (r_g_ shared; an estimate of the concordance of effect direction of shared variants) and genome-wide (r_g_; an estimate of the concordance of effect direction across all genetic variants) were correlated with the prediction of BIP (r_g_ shared: r_*R*_2 =0.90, p=0.03; r_AUC_=0.90, p=0.03; r_g_ genome-wide: r_*R*_2 =0.91, p=0.03; r_AUC_=0.90, p=0.04) but not SCZ (r_g_ shared: r_*R*_2 =0.14, p=0.82; r_AUC_=0.17, p=0.78; r_g_ genome-wide: r_*R*_2 =0.13, p=0.83; r_AUC_=0.16, p=0.80). The Dice coefficient, an estimate of the degree of non-correlative overlap between primary and secondary phenotypes, was not correlated with the prediction of BIP or SCZ (Supplementary Table 3). Similarly, the fraction of SNP-heritability explained by genome-wide significant variants given the current GWAS sample size, an estimate of the power of the GWAS for the secondary phenotype, was not correlated with the prediction of BIP or SCZ (Supplementary Table 3).

**Figure 5.**
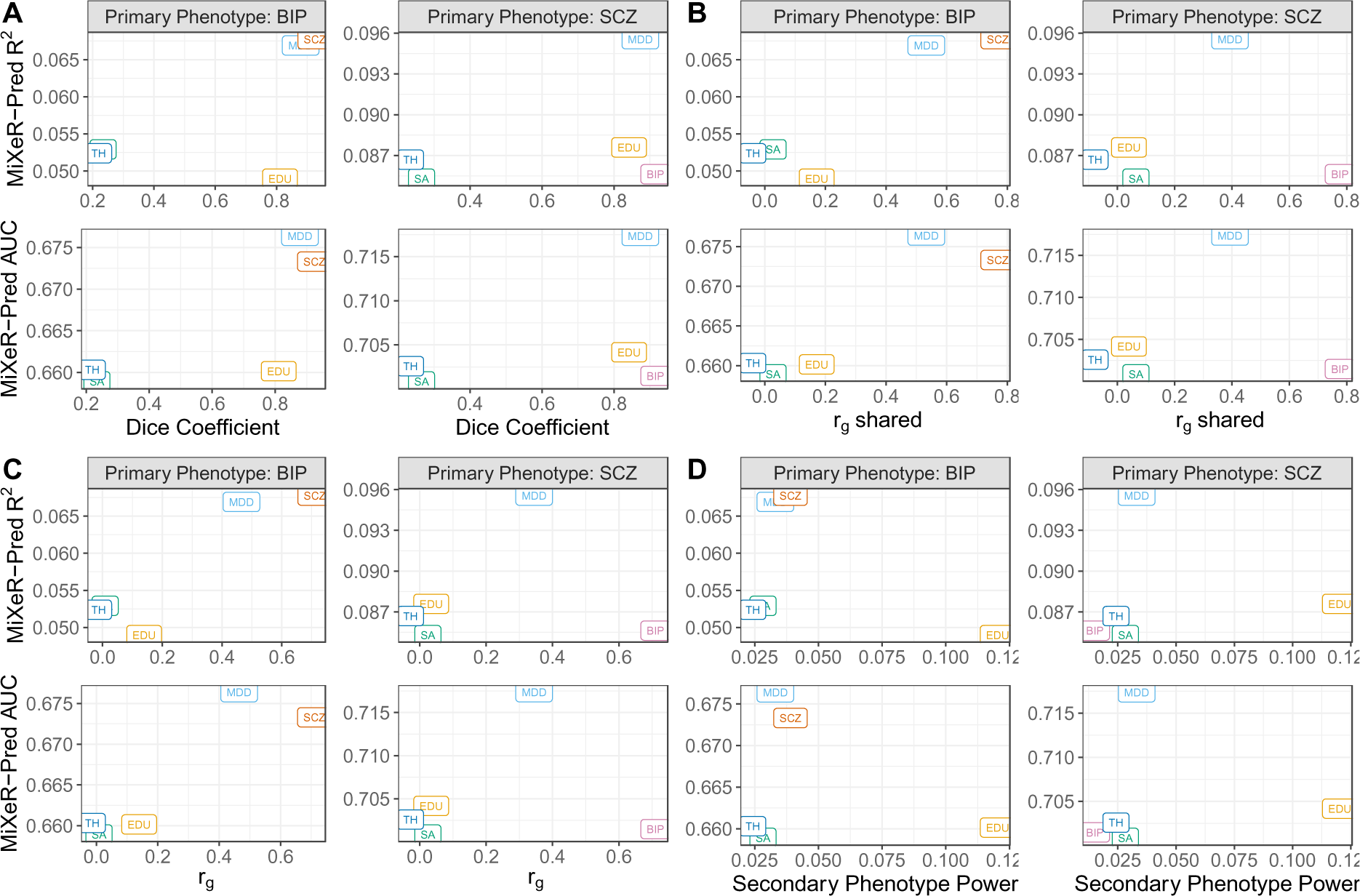
Comparison of MiXeR-Pred Performance and MiXeR Estimates. Performance of MiXeR-Pred measured by variance explained in the primary phenotype (R^2^) and area under the receiver operator curve (AUC) are compared to MiXeR estimates of (A) non-correlative genetic overlap (dice coefficient), (B and C) correlative genetic overlap in the shared component (r_g_ shared) and genome-wide (r_g_), and (D) the current genome-wide association study power of the secondary phenotype. Secondary phenotypes are labelled in boxes within each plot. BIP: bipolar disorder, SCZ: schizophrenia, EDU: education attainment, MDD: major depressive disorder, SA: cortical surface area, TH: cortical thickness.

## Discussion

Here we show that MiXeR-Pred can source genetic overlap to improve prediction performance. We establish that the use of standardized effect estimates as weights for PGS computation, such as Z and IVW, improves the prediction of both BIP and SCZ. Moreover, as MiXeR-Pred parameters are derived from GWAS Z-scores we display the strong association between MiXeR-Pred parameters and standardized effect estimates. For both BIP and SCZ, MiXeR-Pred exhibited greater predictive performance than using the respective univariate GWAS and MTAG, a comparable alternative approach. These findings establish MiXeR-Pred as a novel PGS method that can advance our understanding of pleiotropy between complex traits and disorders while providing foundations for future development of prediction tools that may one day obtain clinical relevance.

There are a large and ever-growing number of PGS methods, few of which use pleiotropy across phenotypes for prediction. The use of multiple polygenic scores has been shown to improve prediction of a primary phenotype of interest but this approach does not produce an improved PGS for the primary phenotype specifically.^17–19^ MTAG is a commonly used method that leverages genetic overlap to boost discovery and prediction.^14^ Here, we demonstrate that MiXeR-Pred had greater predictive performance for BIP and SCZ, two complex psychiatric disorders, using an array of different secondary phenotypes. Consistent with previous reports, both MTAG and MiXeR-Pred, prediction performance was greatest when sourcing genetically correlated secondary phenotypes.^14,40^ A degree of the improved prediction performance observed by MiXeR-Pred is derived from improved modelling of the primary phenotype’s genetic architecture. We also show that for MiXeR-Pred, the prediction from genetically correlated phenotypes remains to some degree when those phenotypes are made to be uncorrelated by randomly shuffling effect directions for the genetic variants. Therefore, MiXeR-Pred can utilize uncorrelated secondary phenotypes for prediction. However, additional prediction performance above univariate factors is likely due to leveraging the phenomenon of mixed effect directions, whereby genetic overlap is characterized by a mixture of concordant and non-concordant effect directions, which may aid in variant selection above and beyond genetic correlation.

MiXeR-Pred performance may be associated with several factors at the univariate and bivariate level. MiXeR-Pred incorporates functional annotations, heterozygosity, and uses a non-infinitesimal approach where genetic variants are considered to have null or non-null effects. These factors combine to improve the modelling of the genetic architecture for a given phenotype and are known to contribute to increased prediction and GWAS replication rates.^8,9,14,31–34,40^ Additionally, MiXeR-Pred uses parameters derived from GWAS Z-scores. We show that using standardized effect estimates as weights (i.e., Z and IVW) can improve prediction above beta coefficients (or log odds ratios). Standardized effect estimates help select more robust effects with smaller estimated error, an approach commonly used for GWAS meta-analysis.^20^ However, as GWAS become more powered and beta coefficients are estimated with reduced error, prediction performance using betas will likely converge with that of standardized effects. We observe indications of this phenomenon as the less powered BIP GWAS has a much larger discrepancy in prediction performance when using beta weights compared to Z and IVW. Conversly, the more powered SCZ GWAS has similar prediction regardless of the weights used. Given a pair of phenotypes, MiXeR-Pred may additionally exploit the degree of polygenic overlap and the power of the secondary phenotype to improve prediction performance. Here we find evidence that correlative genetic overlap contributes to the MiXeR-Pred performance. For example, when predicting BIP, an increase in genetic correlation with the secondary phenotype (i.e., both genome-wide and in the shared component) was associated with an increase in prediction performance. However, this was not the case for SCZ. It is likely that a combination of factors beyond genetic correlation contribute to MiXeR-Pred performance. One such factor is SNP selection with results suggesting this may contribute more to MiXeR-Pred prediction performance than the estimated SNP weights, particularly for genetically uncorrelated secondary phenotypes. This is in line with a previous study which revealed that thresholding variants using false discovery rate values derived from conditioning a GWAS for SCZ on a GWAS for brain morphology improved prediction.^16^ Therefore, a combination of univariate and bivariate factors contribute to the prediction performance of MiXeR-Pred and that combination may differ based on the selection of primary and secondary phenotypes.

MiXeR-Pred has several potential applications. While MiXeR-Pred performance for our test phenotypes remains well below the threshold for clinical utility, future PGS-based approaches can leverage our approach to incorporate both correlated and non-correlated phenotypes to increase predictive performance. MiXeR-Pred expands upon the standard bivariate MiXeR analysis which to date characterizes the shared genetic architecture between two phenotypes beyond genetic correlation.^12^ The addition of MiXeR-Pred therefore enables a more comprehensive interrogation of the genetic overlap between two phenotypes by testing its effect on out-of-sample prediction.

MiXeR-Pred is a conceptually new approach to leveraging pleiotropy for prediction. Therefore, we implement our novel method using the standard pruning and thresholding approach. Other PGS approaches achieve boosts in prediction performance by estimating the direct causal effect of SNPs accounting for LD and avoiding the use of a pruning and thresholding approach.^7–9^ While these approaches perform well for many phenotypes, for some highly polygenic traits they exhibit limited improvement.^6,9^ Future work will expand on the current MiXeR-Pred implementation to further move beyond the standard pruning and thresholding approach. Restricting our analyses to European ancestry only is a limitation. This is because the current implementation of MiXeR relies on a European reference genome. Additionally, the bivariate nature of MiXeR-Pred allows the use of only one secondary phenotype. Future work will expand the MiXeR framework to address these limitations by including multiple ancestries and a tri-variate extension which can ultimately be implemented in MiXeR-Pred.

In conclusion, we present MiXeR-Pred as a novel PGS prediction tool that can source pleiotropy from a secondary phenotype. MiXeR-Pred can use both genetically correlated and to a lesser extent non-correlated secondary phenotypes to improve prediction. The use of our tool can help expand our understanding of the genetic overlap among complex phenotypes. Future work expanding on this framework may ultimately improve our ability to utilize pleiotropy across phenotypes for clinically relevant prediction.

## Data Availability

GWAS summary statistics used in this study are publicly available with the exception of summary statistics excluding the TOP sample which were provided by the psychiatric genomics consortium. The TOP sample data are not publicly available due to national data privacy regulations. The MiXeR-Pred tool presented in this article is available at https://github.com/precimed/mixer-pred. The remaining software are also publicly available: PRSice-2, https://choishingwan.github.io/PRSice/; MTAG, https://github.com/JonJala/mtag; PLINK, https://www.cog-genomics.org/plink/; cleansumstats pipeline used for harmonizing GWAS summary statistics: https://github.com/precimed/python_convert (v0.9.1).

## Acknowledgements

Funding was provided by the Research Council of Norway [grants 223273, 300309, 324252, 326813, 324499, 334920], the South-East Regional Health Authority [grant 2022-073], EEA and Norway [grant EEA-RO-NO-2018-0573], European Union’s Horizon 2020 Research and Innovation Programme [Grant 847776, 964874, 801133], and the National Institutes of Health [grants U24DA041123, U24DA055330].

This work was performed on resources provided by UNINETT Sigma2 (the National Infrastructure for High Performance Computing and Data Storage in Norway) and services from sensitive data (TSD; Tjeneste for Sensitive Data) facilities, University of Oslo Norway.

## Author Contributions

N.P., W.C., and O.F. conducted analyses. N.P., W.C., A.A.S., K.S.O., P.P., E.H., O.A.A., O.F., and A.M.D. were involved in study design and provided analytical input. N.P., G.F.L.H., and O.F. wrote the initial draft of the manuscript. All authors contributed to data interpretation and editing of the manuscript.

## References

1. Visscher, P.M., Wray, N.R., Zhang, Q., Sklar, P., McCarthy, M.I., Brown, M.A., and Yang, J. (2017). 10 Years of GWAS Discovery: Biology, Function, and Translation. The American Journal of Human Genetics 101, 5–22. 10.1016/j.ajhg.2017.06.005.

2. Choi, S.W., Mak, T.S.-H., and O’Reilly, P.F. (2020). Tutorial: a guide to performing polygenic risk score analyses. Nat Protoc 15, 2759–2772. 10.1038/s41596-020-0353-1.

3. Lewis, C.M., and Vassos, E. (2020). Polygenic risk scores: from research tools to clinical instruments. Genome Medicine 12, 44. 10.1186/s13073-020-00742-5.

4. Torkamani, A., Wineinger, N.E., and Topol, E.J. (2018). The personal and clinical utility of polygenic risk scores. Nat Rev Genet 19, 581–590. 10.1038/s41576-018-0018-x.

5. Pain, O., Glanville, K.P., Hagenaars, S.P., Selzam, S., Fürtjes, A.E., Gaspar, H.A., Coleman, J.R.I., Rimfeld, K., Breen, G., Plomin, R., et al. (2021). Evaluation of polygenic prediction methodology within a reference-standardized framework. PLOS Genetics 17, e1009021. 10.1371/journal.pgen.1009021.

6. Ni, G., Zeng, J., Revez, J.A., Wang, Y., Zheng, Z., Ge, T., Restuadi, R., Kiewa, J., Nyholt, D.R., Coleman, J.R.I., et al. (2021). A Comparison of Ten Polygenic Score Methods for Psychiatric Disorders Applied Across Multiple Cohorts. Biol Psychiatry 90, 611–620. 10.1016/j.biopsych.2021.04.018.

7. Privé, F., Arbel, J., and Vilhjálmsson, B.J. (2020). LDpred2: better, faster, stronger. Bioinformatics 36, 5424–5431. 10.1093/bioinformatics/btaa1029.

8. Lloyd-Jones, L.R., Zeng, J., Sidorenko, J., Yengo, L., Moser, G., Kemper, K.E., Wang, H., Zheng, Z., Magi, R., Esko, T., et al. (2019). Improved polygenic prediction by Bayesian multiple regression on summary statistics. Nat Commun 10, 5086. 10.1038/s41467-019-12653-0.

9. Zabad, S., Gravel, S., and Li, Y. (2023). Fast and accurate Bayesian polygenic risk modeling with variational inference. The American Journal of Human Genetics 110, 741–761. 10.1016/j.ajhg.2023.03.009.

10. Watanabe, K., Stringer, S., Frei, O., Umićević Mirkov, M., de Leeuw, C., Polderman, T.J.C., van der Sluis, S., Andreassen, O.A., Neale, B.M., and Posthuma, D. (2019). A global overview of pleiotropy and genetic architecture in complex traits. Nature Genetics 51, 1339–1348. 10.1038/s41588-019-0481-0.

11. Bulik-Sullivan, B.K., Loh, P.-R., Finucane, H.K., Ripke, S., Yang, J., Patterson, N., Daly, M.J., Price, A.L., and Neale, B.M. (2015). LD Score regression distinguishes confounding from polygenicity in genome-wide association studies. Nat Genet 47, 291–295. 10.1038/ng.3211.

12. Frei, O., Holland, D., Smeland, O.B., Shadrin, A.A., Fan, C.C., Maeland, S., O’Connell, K.S., Wang, Y., Djurovic, S., Thompson, W.K., et al. (2019). Bivariate causal mixture model quantifies polygenic overlap between complex traits beyond genetic correlation. Nat Commun 10, 2417. 10.1038/s41467-019-10310-0.

13. Andreassen, O.A., Thompson, W.K., Schork, A.J., Ripke, S., Mattingsdal, M., Kelsoe, J.R., Kendler, K.S., O’Donovan, M.C., Rujescu, D., Werge, T., et al. (2013). Improved Detection of Common Variants Associated with Schizophrenia and Bipolar Disorder Using Pleiotropy-Informed Conditional False Discovery Rate. PLoS Genet 9, e1003455. 10.1371/journal.pgen.1003455.

14. Turley, P., Walters, R.K., Maghzian, O., Okbay, A., Lee, J.J., Fontana, M.A., Nguyen-Viet, T.A., Wedow, R., Zacher, M., Furlotte, N.A., et al. (2018). Multi-trait analysis of genome-wide association summary statistics using MTAG. Nat Genet 50, 229–237. 10.1038/s41588-017-0009-4.

15. Smeland, O.B., Frei, O., Shadrin, A., O’Connell, K., Fan, C.-C., Bahrami, S., Holland, D., Djurovic, S., Thompson, W.K., and Dale, A.M. (2019). Discovery of shared genomic loci using the conditional false discovery rate approach. Human genetics, 1–10.

16. van der Meer, D., Shadrin, A.A., O’Connell, K., Bettella, F., Djurovic, S., Wolfers, T., Alnæs, D., Agartz, I., Smeland, O.B., Melle, I., et al. (2022). Boosting Schizophrenia Genetics by Utilizing Genetic Overlap With Brain Morphology. Biol Psychiatry, S0006–3223(21)01865-5. 10.1016/j.biopsych.2021.12.007.

17. Albiñana, C., Zhu, Z., Schork, A.J., Ingason, A., Aschard, H., Brikell, I., Bulik, C.M., Petersen, L.V., Agerbo, E., Grove, J., et al. (2022). Multi-PGS enhances polygenic prediction: weighting 937 polygenic scores. Preprint at medRxiv, 10.1101/2022.09.14.22279940 10.1101/2022.09.14.22279940.

18. Cheng, W., Parker, N., Karadag, N., Koch, E., Hindley, G., Icick, R., Shadrin, A., O’Connell, K.S., Bjella, T., Bahrami, S., et al. (2023). The relationship between cannabis use, schizophrenia, and bipolar disorder: a genetically informed study. The Lancet Psychiatry 10, 441–451. 10.1016/S2215-0366(23)00143-8.

19. Krapohl, E., Patel, H., Newhouse, S., Curtis, C.J., von Stumm, S., Dale, P.S., Zabaneh, D., Breen, G., O’Reilly, P.F., and Plomin, R. (2018). Multi-polygenic score approach to trait prediction. Mol Psychiatry 23, 1368–1374. 10.1038/mp.2017.163.

20. Willer, C.J., Li, Y., and Abecasis, G.R. (2010). METAL: fast and efficient meta-analysis of genomewide association scans. Bioinformatics 26, 2190–2191. 10.1093/bioinformatics/btq340.

21. Hindley, G., Frei, O., Shadrin, A.A., Cheng, W., O’Connell, K.S., Icick, R., Parker, N., Bahrami, S., Karadag, N., Roelfs, D., et al. (2022). Charting the Landscape of Genetic Overlap Between Mental Disorders and Related Traits Beyond Genetic Correlation. AJP, appi.ajp.21101051. 10.1176/appi.ajp.21101051.

22. Mullins, N., Forstner, A.J., O’Connell, K.S., Coombes, B., Coleman, J.R.I., Qiao, Z., Als, T.D., Bigdeli, T.B., Børte, S., Bryois, J., et al. (2021). Genome-wide association study of more than 40,000 bipolar disorder cases provides new insights into the underlying biology. Nat Genet 53, 817–829. 10.1038/s41588-021-00857-4.

23. Trubetskoy, V., Pardiñas, A.F., Qi, T., Panagiotaropoulou, G., Awasthi, S., Bigdeli, T.B., Bryois, J., Chen, C.-Y., Dennison, C.A., Hall, L.S., et al. (2022). Mapping genomic loci implicates genes and synaptic biology in schizophrenia. Nature 604, 502–508. 10.1038/s41586-022-04434-5.

24. Bansal, V., Mitjans, M., Burik, C. a. P., Linnér, R.K., Okbay, A., Rietveld, C.A., Begemann, M., Bonn, S., Ripke, S., de Vlaming, R., et al. (2018). Genome-wide association study results for educational attainment aid in identifying genetic heterogeneity of schizophrenia. Nat Commun 9, 3078. 10.1038/s41467-018-05510-z.

25. Comes, A.L., Senner, F., Budde, M., Adorjan, K., Anderson-Schmidt, H., Andlauer, T.F.M., Gade, K., Hake, M., Heilbronner, U., Kalman, J.L., et al. (2019). The genetic relationship between educational attainment and cognitive performance in major psychiatric disorders. Transl Psychiatry 9, 1–11. 10.1038/s41398-019-0547-x.

26. Grasby, K.L., and Jahanshad, N. (2020). The genetic architecture of the human cerebral cortex. Science, 17.

27. Smeland, O.B., Kutrolli, G., Bahrami, S., Fominykh, V., Parker, N., Hindley, G.F.L., Rødevand, L., Jaholkowski, P., Tesfaye, M., Parekh, P., et al. (2023). The shared genetic risk architecture of neurological and psychiatric disorders: a genome-wide analysis. Preprint at medRxiv, 10.1101/2023.07.21.23292993 10.1101/2023.07.21.23292993.

28. Cheng, W., Frei, O., van der Meer, D., Wang, Y., O’Connell, K.S., Chu, Y., Bahrami, S., Shadrin, A.A., Alnæs, D., Hindley, G.F.L., et al. (2021). Genetic Association Between Schizophrenia and Cortical Brain Surface Area and Thickness. JAMA Psychiatry 78, 1020–1030. 10.1001/jamapsychiatry.2021.1435.

29. Cheng, W., van der Meer, D., Parker, N., Hindley, G., O’Connell, K.S., Wang, Y., Shadrin, A.A., Alnæs, D., Bahrami, S., Lin, A., et al. (2022). Shared genetic architecture between schizophrenia and subcortical brain volumes implicates early neurodevelopmental processes and brain development in childhood. Mol Psychiatry, 1–10. 10.1038/s41380-022-01751-z.

30. Parker, N., Cheng, W., Hindley, G.F.L., Parekh, P., Shadrin, A.A., Maximov, I.I., Smeland, O.B., Djurovic, S., Dale, A.M., Westlye, L.T., et al. (2023). Psychiatric disorders and brain white matter exhibit genetic overlap implicating developmental and neural cell biology. Mol Psychiatry, 1–9. 10.1038/s41380-023-02264-z.

31. Frei, O., Hindley, G., Shadrin, A.A., Meer, D. van der Akdeniz, B.C., Cheng, W., O’Connell, K.S., Bahrami, S., Parker, N., Smeland, O.B., et al. (2022). Improved functional mapping with GSA-MiXeR implicates biologically specific gene-sets and estimates enrichment magnitude. Preprint at medRxiv, 10.1101/2022.12.08.22283159 10.1101/2022.12.08.22283159.

32. Zheng, Z., Liu, S., Sidorenko, J., Yengo, L., Turley, P., Wang, R., Nolte, I.M., Snieder, H., Wray, N.R., Goddard, M.E., et al. Leveraging functional genomic annotations and genome coverage to improve polygenic prediction of complex traits within and between ancestries.

33. Márquez-Luna, C., Gazal, S., Loh, P.-R., Kim, S.S., Furlotte, N., Auton, A., and Price, A.L. (2021). Incorporating functional priors improves polygenic prediction accuracy in UK Biobank and 23andMe data sets. Nat Commun 12, 6052. 10.1038/s41467-021-25171-9.

34. Wang, Y., Thompson, W.K., Schork, A.J., Holland, D., Chen, C.-H., Bettella, F., Desikan, R.S., Li, W., Witoelar, A., Zuber, V., et al. (2016). Leveraging Genomic Annotations and Pleiotropic Enrichment for Improved Replication Rates in Schizophrenia GWAS. PLOS Genetics 12, e1005803. 10.1371/journal.pgen.1005803.

35. Chang, C.C., Chow, C.C., Tellier, L.C., Vattikuti, S., Purcell, S.M., and Lee, J.J. (2015). Second-generation PLINK: rising to the challenge of larger and richer datasets. GigaScience 4, s13742–015-0047–0048. 10.1186/s13742-015-0047-8.

36. Choi, S.W., and O’Reilly, P.F. (2019). PRSice-2: Polygenic Risk Score software for biobank-scale data. GigaScience 8, giz082. 10.1093/gigascience/giz082.

37. Okbay, A., Wu, Y., Wang, N., Jayashankar, H., Bennett, M., Nehzati, S.M., Sidorenko, J., Kweon, H., Goldman, G., Gjorgjieva, T., et al. (2022). Polygenic prediction of educational attainment within and between families from genome-wide association analyses in 3 million individuals. Nat Genet 54, 437–449. 10.1038/s41588-022-01016-z.

38. Als, T.D., Kurki, M.I., Grove, J., Voloudakis, G., Therrien, K., Tasanko, E., Nielsen, T.T., Naamanka, J., Veerapen, K., Levey, D.F., et al. (2023). Depression pathophysiology, risk prediction of recurrence and comorbid psychiatric disorders using genome-wide analyses. Nat Med 29, 1832–1844. 10.1038/s41591-023-02352-1.

39. Lee, S.H., Goddard, M.E., Wray, N.R., and Visscher, P.M. (2012). A Better Coefficient of Determination for Genetic Profile Analysis. Genetic Epidemiology 36, 214–224. 10.1002/gepi.21614.

40. Hu, Y., Lu, Q., Liu, W., Zhang, Y., Li, M., and Zhao, H. (2017). Joint modeling of genetically correlated diseases and functional annotations increases accuracy of polygenic risk prediction. PLOS Genetics 13, e1006836. 10.1371/journal.pgen.1006836.

