## Supplementary Methods for "Sourcing Bivariate Genetic Overlap for Polygenic Prediction using MiXeR-Pred"

#### The Norwegian Thematically Organized Psychosis Sample

We conduct our polygenic score (PGS) analysis using the Norwegian Thematically Organized Psychosis (TOP) sample. The TOP sample includes participants recruited from regions across Norway [Oslo, Trondheim, and several Southeast regional hospitals (Diakonhjemmet Hospital, Lovisenberg Hospital, and St Olav's Hospital)]. DSM-IV criteria were used for the inclusion of diagnosed BIP and SCZ participants. Healthy controls, with a similar age to the cases, were randomly selected from the same catchment area as the cases. Participants provided written consent and the study was approved by the Regional Committee for Medical and Health Research Ethics of South-East Norway.

PLINK v1.9 was used to perform SNP and sample quality control. SNPs were filtered using thresholds for call rates (5% missingness), minor allele frequencies (0.01), and departure from Hardy-Weinberg equilibrium (1E-06). Samples with missingness above 2% were removed.

#### Clumpling Procedure

MiXeR-Pred uses PLINK v1.9 to clump SNPs based on the derived thresholding value ($\exp(-E\delta_1^2)$). We used the 1000 genomes European sample for clumping. Below are the default PLINK commands:

plink --bfile 1000G_EUR --clump-p1 1 --clump-r2 0.2 --clump-kb 250 --clump "MiXeR_Pred_file" --climp-snp-field RSID --clump-field "MiXeR_Pred_threshold" –out "output_filename"

Note: for MTAG the "mtag_pval" values were used for clumping while for the standard GWAS clumping we use the p-value provided in the summary statistics.

#### Previous Implementations of Univariate and Bivariate MiXeR

MiXeR v1.3[1,2] was used here for estimation of several parameters in the analyses of contributions to MiXeR-Pred prediction. These parameters include the Dice coefficient, genome-wide genetic correlation ($r_g$), genitive correlation within the shared/overlapping component ($r_g$ shared), and the explained variance in heritability given the current sample size of the GWAS (power). Details on MiXeR v1.3 implementation are described below.

For each SNP, i, MiXeR v1.3 models its additive genetic effect of allele substitution, $\beta_i$, as a point-normal mixture, $\beta_i = (1 - \pi_1)N(0,0) + \pi_1 N(0, \sigma_\beta^2)$, where $\pi_1$ represents the proportion of non-null SNPs (i.e., polygenicity) and $\sigma_\beta^2$ represents the variance of effect sizes of non-null SNPs (i.e., discoverability). For each SNP, j, MiXeR v1.3 incorporates LD information and allele frequencies for background SNPs extracted from 1000 Genomes Phase3 data to estimate the expected probability distribution of the signed test statistic, $z_j = \delta_j + \epsilon_j = N \sum_i \sqrt{H_i} r_{ij} \beta_i + \epsilon_j$, where $N$ is the sample size, $H_i$ indicates heterozygosity of i-th SNP, $r_{ij}$ indicates allelic correlation between i-th and j-th SNPs, and $\epsilon_j \sim N(0, \sigma_0^2)$ is the residual variance. Further, the three parameters, $\pi_1, \sigma_\beta^2, \sigma_0^2$, are fitted by direct maximization of the likelihood function. The number of independent SNPs is estimated as $M\pi_1$, where M is the number of selected background SNPs in the reference panel. Phenotypic variance explained on average by an independent SNP is calculated as $\overline{H}\sigma_\beta^2$, where $\overline{H} = \frac{1}{M}\sum_i H_i = 0.2075$ is the average heterozygosity across SNPs in the reference panel. Under the assumptions of the MiXeR model, SNP-heritability is then calculated as $h_{SNP}^2 = M\pi_1 \times \overline{H}\sigma_\beta^2$.

The bivariate analysis models additive genetic effects as a mixture of four components: 1) SNPs that are null in both phenotypes ($\pi_0$), (2) SNPs with an effect on the first phenotype $\pi_1$, (3) SNPs with an effect on the second phenotypes ($\pi_2$), and (4) SNPs with an effect on both phenotypes ($\pi_{12}$). In the last component, MiXeR models the variance-covariance matrix as $\boldsymbol{\Sigma_{12}} = \begin{bmatrix} \sigma_1^2 & \rho_{12}\sigma_1\sigma_2 \\ \rho_{12}\sigma_1\sigma_2 & \sigma_2^2 \end{bmatrix}$ where $\rho_{12}$ indicates correlation of effect sizes within the shared component, and $\sigma_1^2$ and $\sigma_2^2$ correspond to the discoverability parameter estimated in the univariate analysis of the two tr phenotypes. After fitting parameters of the model genetic correlation is calculated as $r_g = \frac{\rho_{12}\pi_{12}}{\sqrt{(\pi_1+\pi_{12})(\pi_2+\pi_{12})}}$. The Dice coefficient (DC) was calculated using the formula DC = $\frac{2\pi_{12}}{\pi_1+\pi_2+2\pi_{12}}$.

To determine the suitability of summary statistics and the sufficiency of model fit, MiXeR v1.3 employs the Akaike information criterion ($AIC = 2k - 2\ln L$ ), where $k$ is the number of free parameters in the model, $L$ is the value of the likelihood function, and $n$ is the effective number of SNPs used in the optimization procedure. The AIC represents the comparison between MiXeR v1.3 modelled fit and the infinitesimal model, which assumes an all SNPs influence a phenotype with some having an infinitesimal effect. A positive AIC value indicates the input GWAS dataset provides sufficient power to discriminate the MiXeR v1.3 modelled fit from infinitesimal model and the MiXeR v1.3 estimates are reliable.

#### PRSice2 Estimation of Variance Explained by the Polygenic Score

The PRSice2 software calculates the variance explained for binary outcomes, such as case-control status, using the Nagelkerke $R^2$.[3] Two models are used to estimate the $R^2$ for a given polygenic score (PGS). The $R^2$ from a base model containing only covariates is subtracted from the $R^2$ of a full model containing the PGS and covariates. Thus, the PGS $R^2$ is a result of this difference in $R^2$ between the full and base model.

#### Evaluating Contributions of MiXeR-Pred SNP Weights vs Selection

We also assessed the separate contributions of MiXeR-Pred weights and thresholds for SNP selection. For the SNP weights only scenario, we use MiXeR-Pred weights but prune and select top independent SNPs for PGS computation using the primary phenotypes GWAS p-values. For the SNP selection only scenario, we use the primary phenotypes GWAS Z-score as weights but MiXeR-Pred thresholding values for pruning and selection of top independent SNPs for PGS computation. Prediction performance for both the SNP weights and selection scenarios are compared to the approach in the main text which uses both MiXeR-Pred values to generate PGS.

### References

1. Frei, O., Holland, D., Smeland, O.B., Shadrin, A.A., Fan, C.C., Maeland, S., O'Connell, K.S., Wang, Y., Djurovic, S., Thompson, W.K., et al. (2019). Bivariate causal mixture model quantifies polygenic overlap between complex traits beyond genetic correlation. Nat Commun *10*, 2417. 10.1038/s41467-019-10310-0.

2. Holland, D., Frei, O., Desikan, R., Fan, C.-C., Shadrin, A.A., Smeland, O.B., Sundar, V.S., Thompson, P., Andreassen, O.A., and Dale, A.M. (2020). Beyond SNP heritability: Polygenicity and discoverability of phenotypes estimated with a univariate Gaussian mixture model. PLoS Genet *16*, e1008612. 10.1371/journal.pgen.1008612.

3. Euesden, J., Lewis, C.M., and O'Reilly, P.F. (2015). PRSice: Polygenic Risk Score software. Bioinformatics *31*, 1466–1468. 10.1093/bioinformatics/btu848.
