## Supplementary Figures for "Sourcing Bivariate Genetic Overlap for Polygenic Prediction using MiXeR-Pred"

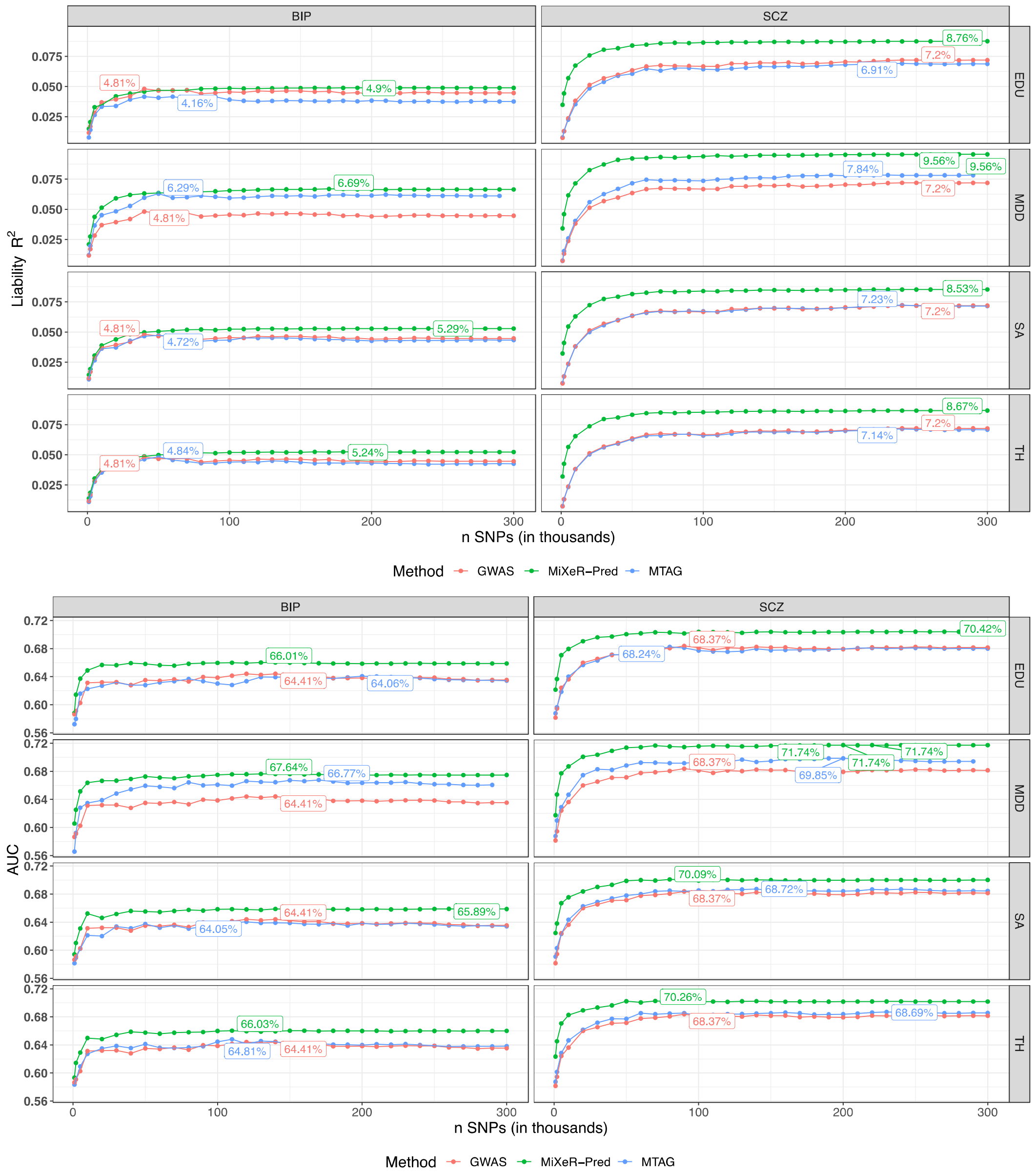


**Supplementary Figure 1. Prediction Comparison Across Methods and Secondary Phenotypes.** Figures compare the MiXeR-Pred polygenic score prediction performance [variance explained ($R^2$) on the liability scale and area under the receiver operator curve (AUC)] to the multi-trait analysis of genome wide association studies (MTAG) and the primary trait's genome wide association study (GWAS) polygenic score (PGS). Education attainment (EDU), major depressive disorder (MDD), cortical surface area (SA), and cortical thickness (TH) are used as secondary phenotypes for bipolar disorder (BIP) and schizophrenia (SCZ). Note that the secondary phenotypes are used for both MiXeR-Pred and MTAG approaches. However, the GWAS PGS performance is constant regardless of secondary phenotype since it is the PGS derived from the primary phenotypes GWAS only.

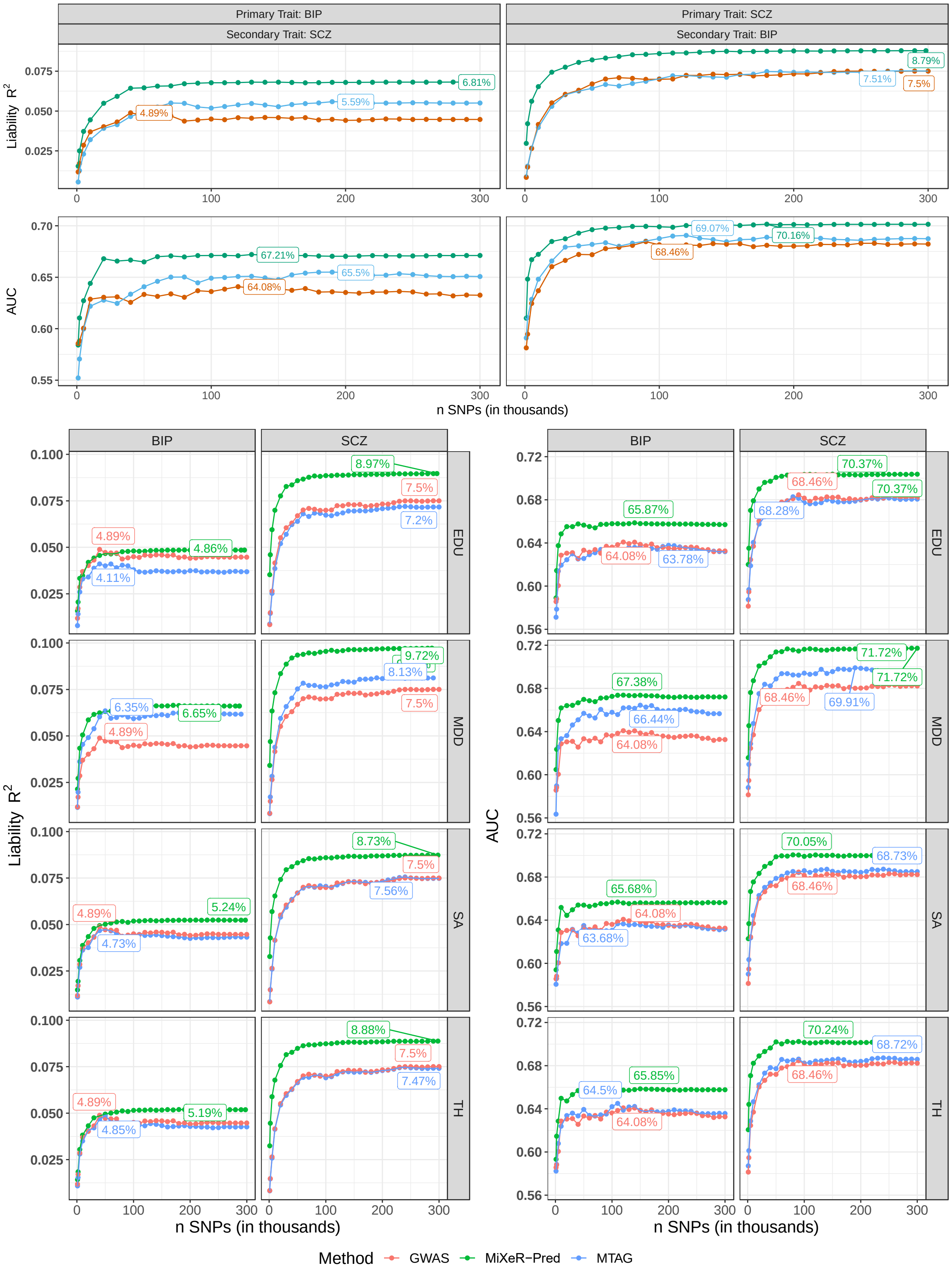


**Supplementary Figure 2. Prediction Comparison Across Methods and Secondary Phenotypes.** Here all models adjust for age, sex, genetic batch, and the first 20 genetic principle components. Figures compare the MiXeR-Pred polygenic score (PGS) prediction performance [variance explained ($R^2$) on the liability scale and area under the receiver operator curve (AUC)] to the multi-trait analysis of genome wide association studies (MTAG) and the primary trait's genome wide association study (GWAS) PGS. First bipolar disorder (BIP) and schizophrenia (SCZ) serve as primary and secondary traits for each other. Education attainment (EDU), major depressive disorder (MDD), cortical surface area (SA), and cortical thickness (TH) are used as secondary phenotypes for BIP and SCZ. Note that the secondary phenotypes are used for both MiXeR-Pred and MTAG approaches. However, the GWAS PGS performance is constant regardless of secondary phenotype since it is the PGS derived from the primary phenotypes GWAS only.

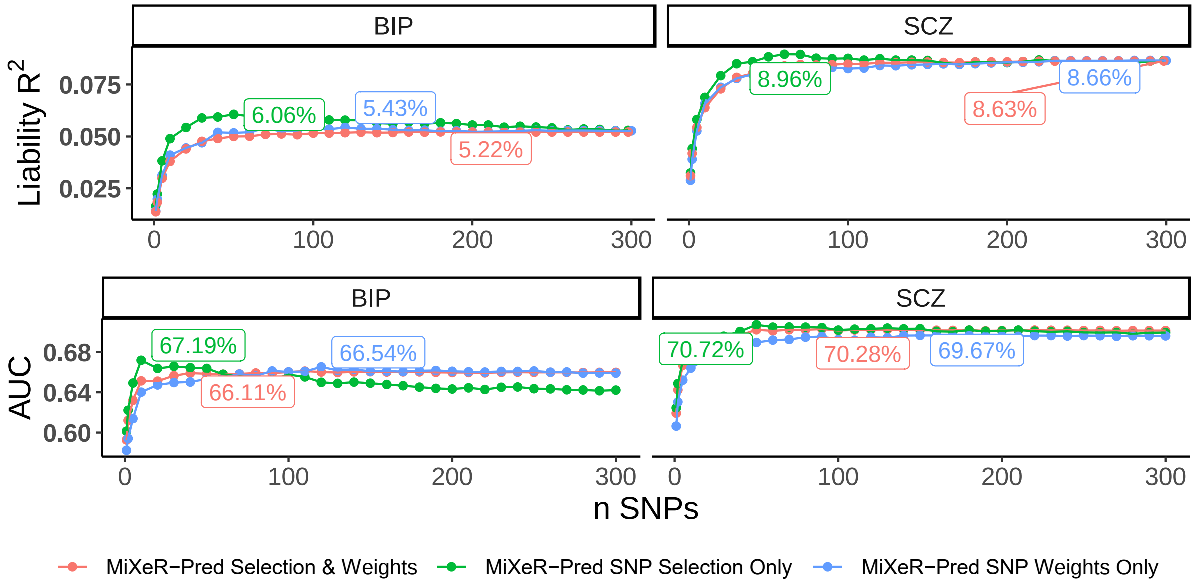


**Supplementary Figure 3. Univariate MiXeR-Pred Prediction Performance.** Here all models adjust for genetic batch and the first 20 genetic principle components. Figures compare the use of univariate MiXeR-Pred weights only (blue), thresholds for variant selection only (green), and both (pink). Polygenic score prediction performance is measured using variance explained ($R^2$) on the liability scale and area under the receiver operator curve (AUC).

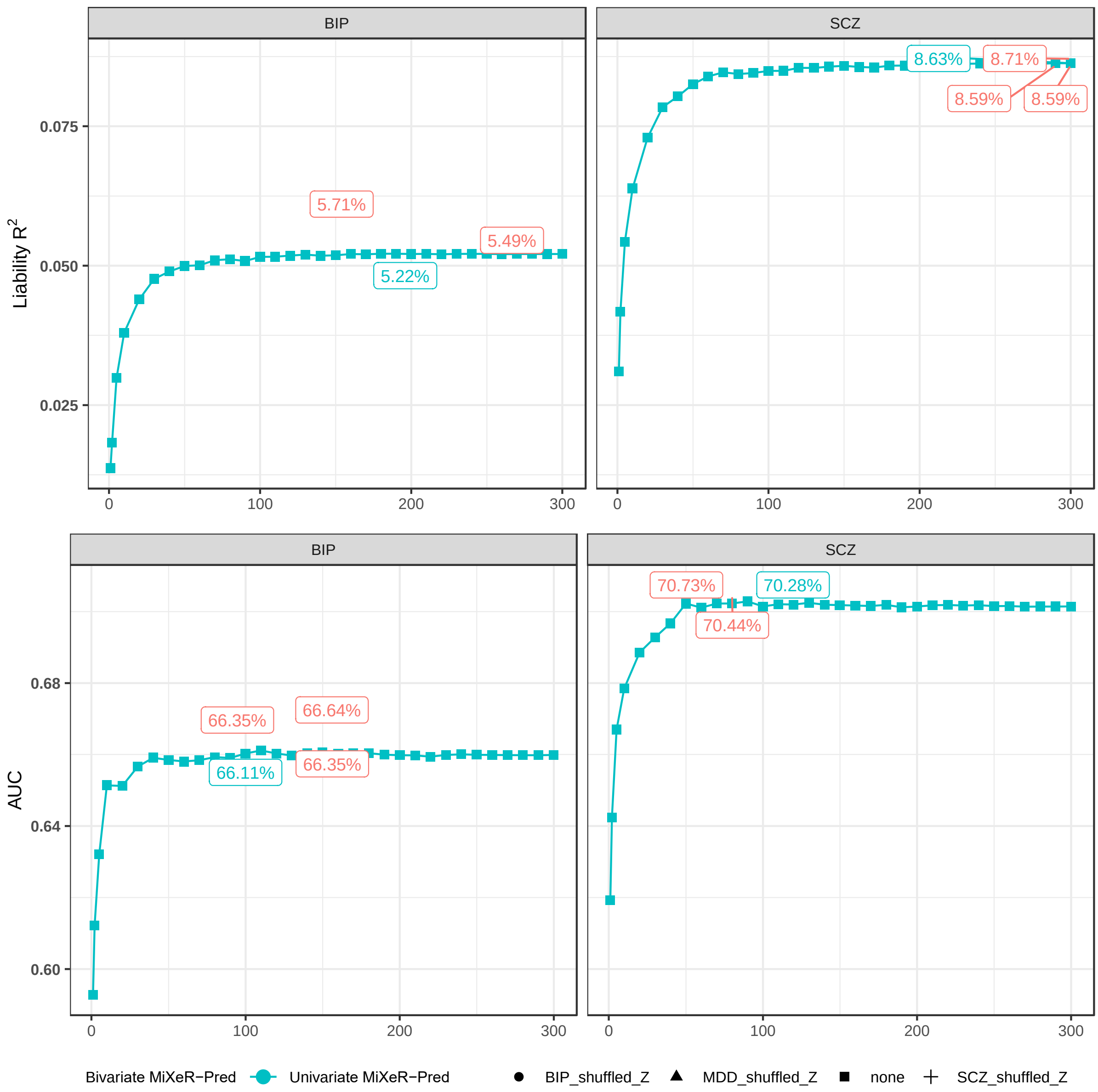


**Supplementary Figure 4. MiXeR-Pred Prediction Performance with Uncorrelated Secondary Phenotypes.** For bipolar disorder (BIP) as a primary phenotype (left), effect directions for major depressive disorder (MDD, triangles) and schizophrenia (SCZ, cross) were shuffled (i.e., MDD_shuffled_Z, SCZ_shuffled_Z) to produce no genetic correlation. For SCZ as a primary phenotype, effect directions of BIP (i.e., BIP_shuffled_Z, circles) MDD were shuffled to produce no genetic correlation. Here all models adjust for genetic batch and the first 20 genetic principle components. Figures compare the use of univariate MiXeR-Pred (green) and bivariate MiXeR-Pred (pink). Polygenic score prediction performance is measured using variance explained ($R^2$) on the liability scale and area under the receiver operator curve (AUC).

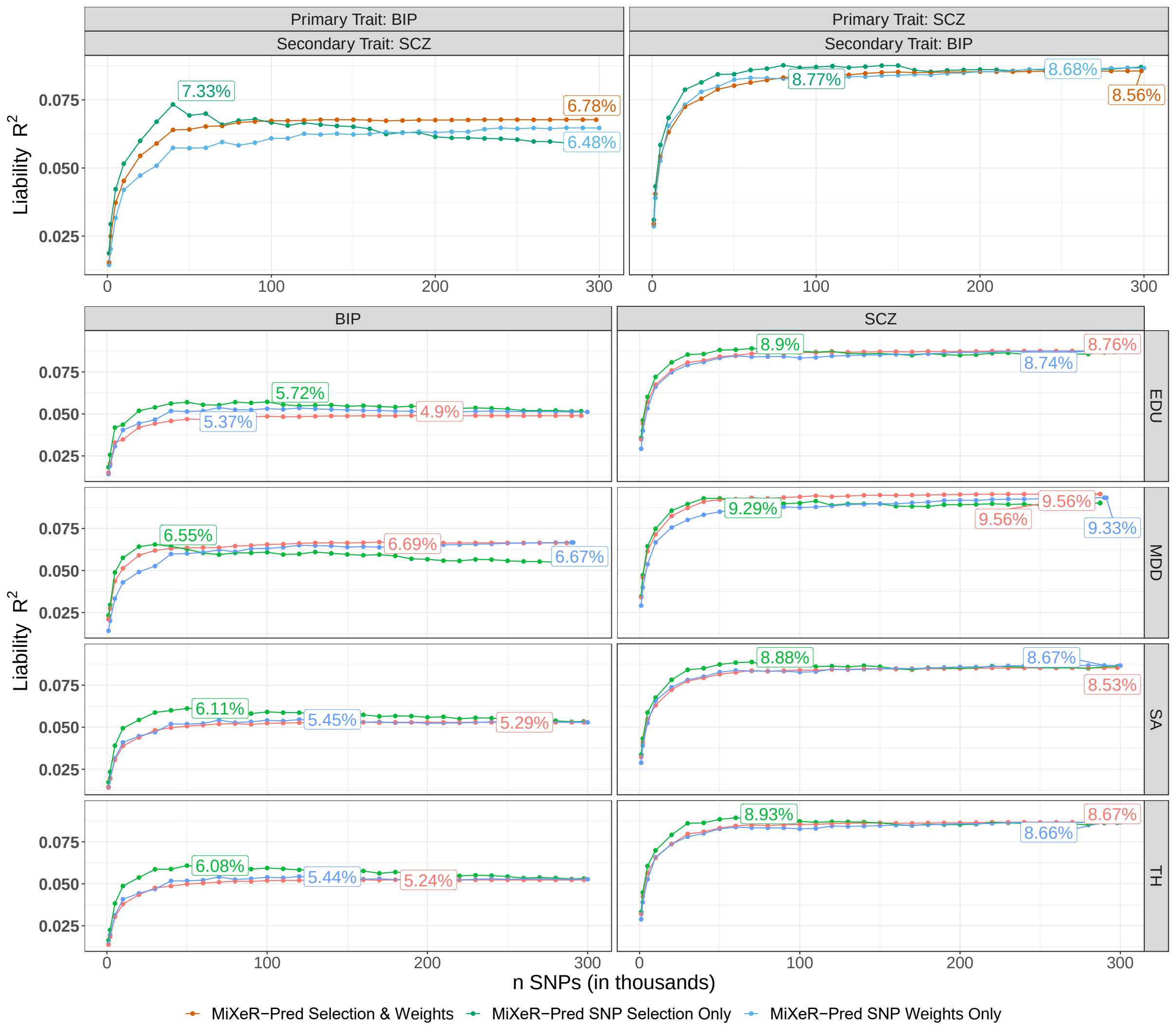


**Supplementary Figure 5. MiXeR-Pred $R^2$ for Polygenic Scores with Weights, Thresholds, or Both.** Here all models adjust for genetic batch and the first 20 genetic principle components. Figures compare the use of MiXeR-Pred weights only (blue), thresholds for variant selection only (green), and both (pink). Polygenic score prediction performance is measured using variance explained ($R^2$) on the liability scale. First bipolar disorder (BIP) and schizophrenia (SCZ) serve as primary and secondary traits for each other. Then education attainment (EDU), major depressive disorder (MDD), cortical surface area (SA), and cortical thickness (TH) are used as secondary phenotypes for BIP and SCZ. Note that the primary phenotypes Z value is used as weights for the selection only scenario while the p-values are used for pruning and thresholding in the weights only scenario.

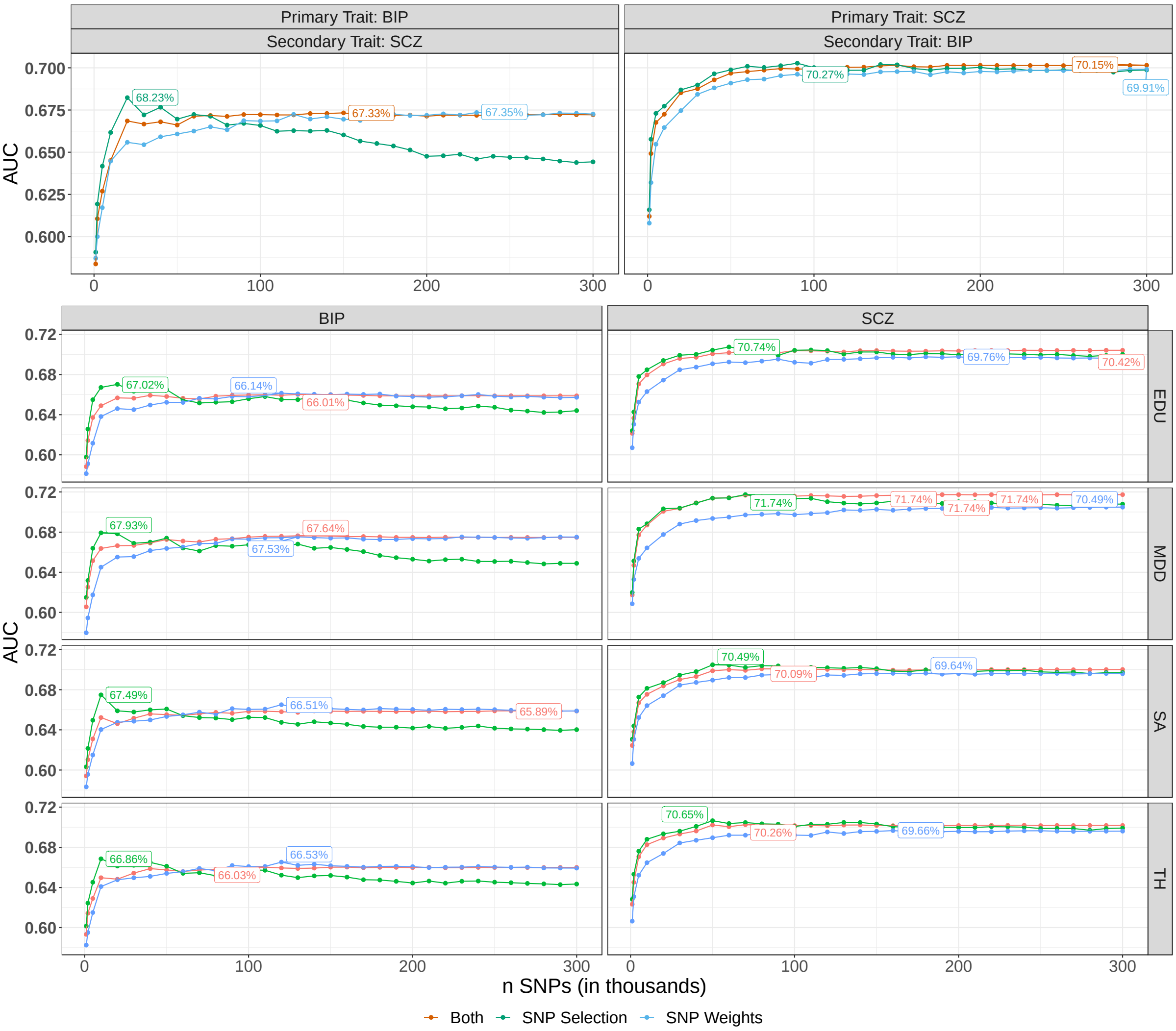


**Supplementary Figure 6. MiXeR-Pred AUC for Polygenic Scores with Weights, Thresholds, or Both.** Here all models adjust for genetic batch and the first 20 genetic principle components. Figures compare the use of MiXeR-Pred weights only (blue), thresholds only for variant selection only (green), and both weights and thresholds (pink). Polygenic score prediction performance is measured using area under the receiver operator curve (AUC). First bipolar disorder (BIP) and schizophrenia (SCZ) serve as primary and secondary traits for each other. Then education attainment (EDU), major depressive disorder (MDD), cortical surface area (SA), and cortical thickness (TH) are used as secondary phenotypes for BIP and SCZ. Note that the primary phenotypes Z value is used as weights for the selection only scenario while the p-values are used for pruning and thresholding in the weights only scenario.
